# Characterization of perceptual and electrophysiological responses to high-speed robot-controlled pinprick stimulation

**DOI:** 10.1101/2025.02.20.25322639

**Authors:** Sofía A. Poux, Abed Youssif, Albano Peñalva, Emanuel N. Van Den Broeke, Ulf Baumgärtner, José A. Biurrun Manresa, Christian A. Mista

## Abstract

**Objective:** This work utilises a novel high-speed robot-controlled stimulator to evaluate the effects of different levels of mechanical impulse (force and duration) of pinprick stimuli on neurophysiological responses and subjective perception.

**Methods:** Twenty-seven healthy volunteers received pinprick stimuli on the right volar forearm using different combinations of force and duration that configured three distinct levels: low, medium and high mechanical impulse stimuli. The robotic system recorded stimulus force and duration in real time, and participants rated the pinprick perceived intensity on a numerical rating scale (NRS). The peak amplitudes and latencies of the pinprick evoked potentials (PEPs) were compared across stimulation levels. Perceived intensity ratings were analysed using linear mixed models, and the PEP features were evaluated through spatio-temporal cluster permutation tests.

**Results:** Perceived intensity ratings, expressed as median [IQR], were 2.5 [1.14 *–* 8.15] for low, 16.4 [ 8.86 *–* 26.9] for medium and 28.5 [10.2 *–* 41.4] for high impulse stimuli. Differences in brain activity were found when comparing across stimulation levels: the N2 Peak latency was longer for high impulse compared to low impulse and for medium impulse compared to low impulse. The P2 peak latency showed no differences across levels. The N2 amplitude was larger for high vs. low impulse and for medium vs. low impulse, with no significant difference observed between high and medium impulse stimulation.

**Conclusions:** High speed robot-controlled pinprick stimulation successfully elicited pinprick-evoked potentials (PEPs) across different mechanical impulse levels. The use of robot-controlled systems may help standardize the assessment of mechanical nociception.

## Introduction

Several pathological conditions result in small fibre neuropathy (SFN), which affects both peripheral nerve fibres with thin myelin sheaths (Aδ fibres) and those without myelin (C fibres) (Pál et al., 2020). To diagnose SFN, clinicians examine a range of clinical signs, including decreased pinprick sensation (Devigili et al., 2020). The assessment of responses of Aδ fibre to mechanical stimulation represents a valuable tool for evaluating sensory pathways. Perceptual responses are usually quantified using numerical rating scales (NRS) and questionnaires, whereas brain responses can be assessed through functional magnetic resonance imaging (fMRI) or electroencephalography (EEG). In particular, electrophysiological responses to time-locked mechanical stimuli are known as pinprick evoked potentials (PEPs). These potentials are transmitted through the spinothalamic pathways and serve as a tool for assessing the excitability of the nociceptive system in experimental and clinical conditions (Madsen et al., 2014).

The first reports on PEPs employed handheld pinprick devices with calibrated weights and electronic trigger systems to detect the contact of the needle with the skin. The first study tested a single pinprick exerting a force of 128 mN (Iannetti et al., 2013). Later, van den Broeke et al. (2015) expanded the range of tested intensities, applying forces from 16 to 512 mN in multiples of two, following the existing sets of pinpricks routinely used for quantitative sensory tests (Baumgärtner et al., 2002; Chan et al., 1992; Rolke et al., 2006). Van den Broeke et al. (2015) and subsequent studies showed that the use of the 64-mN pinprick demonstrated good sensitivity to show changes due to experimental interventions modulating the excitability of the nociceptive system (van den Broeke et al., 2015, 2017, 2019). Moreover, recent studies used forces of 128 mN and 256 mN, to investigate PEP reliability (Rosner et al., 2020; Triccas et al., 2022).

Furthermore, the optimal stimulus duration for eliciting PEPs has not been thoroughly investigated. While some studies have designed their experimental paradigms to apply the stimulus as rapidly as possible (Iannetti et al., 2013), others opted for a sustained stimulation of approximately 1-s duration to prevent inertial effects (Rosner et al., 2020; van den Broeke et al., 2015, 2016, 2017). Recent studies employed robotic systems (Gousset et al., 2023; Jaltare, Manresa, et al., 2024; Jaltare, Meyers, et al., 2024; van den Broeke et al., 2019, 2020), avoiding the manual application of the stimulus and thus reducing the variability of the force applied (Leone et al., 2025; Valentini & Schulz, 2020). Although stimulus parameters were conservatively maintained (same weights and duration), it was suggested that further adjustments are still required (van den Broeke et al., 2020). However, there is scarce information on how the interaction between force and duration, i.e. the mechanical impulse, impacts the response to pinprick stimulation, which might be relevant to untangle the contribution of nociceptive and non-nociceptive fibres to PEPs (Nagi et al., 2019). Thus, the objective of the study is to characterize the perceptual and electrophysiological responses to pinprick stimuli with different mechanical impulse stimulation levels using a high-speed robot-controlled stimulator.

## Materials and Methods

### Study design

Study preregistration, including original hypothesis and description of primary and secondary outcomes, are available at ClinicalTrials.gov (identifier: <u>NCT06183593</u>). A single-group, crossover design was employed in this descriptive study, in which participants were presented with successive trials of pinprick using three stimulation levels (low, medium, and high mechanical impulse), in random order. Written informed consent was obtained from all volunteers before participation. All experimental procedures were approved by the local ethics committee (approval number IS004486) and were conducted according to the principles of the Declaration of Helsinki.

### Sample size considerations

Since this is a descriptive study, sample size was estimated in terms of precision for the main outcome, which was the latency of the N2 peak of the PEPs. Previous studies reported that pinprick stimuli applied to the fingertip with a force of 256 mN resulted in N2 latencies of 89 *±* 25.8 ms (mean and standard deviation) (Rosner et al, 2020). Based on these values, at least 25 subjects are required to obtain a precision of 10 ms (i.e., the width of the 95% confidence intervals around the mean) in the estimation of the N2 peak.

### Participants

Twenty-seven healthy volunteers (11 females) aged between 20 and 52 participated in the experiment. Exclusion criteria were pregnancy, history of chronic pain or neuromuscular disorders, or any disease affecting the nociceptive system that would prevent normal evaluation. The use of analgesics or excessive alcohol intake within 24 hours before the experiment and lack of sleep (less than 6 hours) the night before the experiment, were also considered as exclusion criteria.

### Pinprick stimulation

A custom-made robot-controlled mechanical stimulator was used to deliver the pinprick stimuli. The stimulator consists of a linear actuator (EPCO-16-50-3P, Festo SE & Co. KG, Germany) solidary to a stainless-steel spring (stiffness *k _sp_*=49N/m), a stainless-steel blunt tip (diameter *d*=0.36 mm), and a load cell to measure the applied force in real time during stimulation. The force recording was used to detect the moment when the probe makes contact with the skin, enabling synchronization with the EEG recordings. The magnitude of the applied force during stimulation is directly proportional to the travel distance of the tip and the combined stiffness of the spring (*k _sp_*) and skin (*k _sk_*).

Different combinations of force and duration provide different levels of mechanical impulse, defined as *I* =∫ *F* (*t*) *dt*. With this setup, we tested three different stimulation levels: low, medium and high impulse (Fig. 1). For all these levels, the travel distance of the linear actuator was set to 30 mm with a speed of 125 mm/s. Low impulse stimulation was achieved by positioning the tip 25 mm away from the skin of the volar forearm, moving the tip towards the skin at its maximum speed and removing the tip immediately after the predefined travel distance was reached. Medium impulse stimulation was achieved by positioning the tip 10 mm away from the skin, moving the tip towards the skin at its maximum speed and removing the tip immediately after the predefined travel distance was reached. High impulse stimulation was achieved by positioning the tip 10 mm away from the skin, moving the tip towards the skin at its maximum speed until the predefined travel distance was reached, maintaining the force on the skin for 1 s, and then removing the tip. In all cases, the distance covered by the tip matches the spring compression (shortening) plus skin indentation. In this way, the applied force increased linearly due to the spring’s mechanical properties. Stimulation was repeated 40 times for each stimulation level, with an inter-stimulus interval ranging from 12 to 18 s. The order of application for each stimulation level was randomized across volunteers.

**Figure 1:**
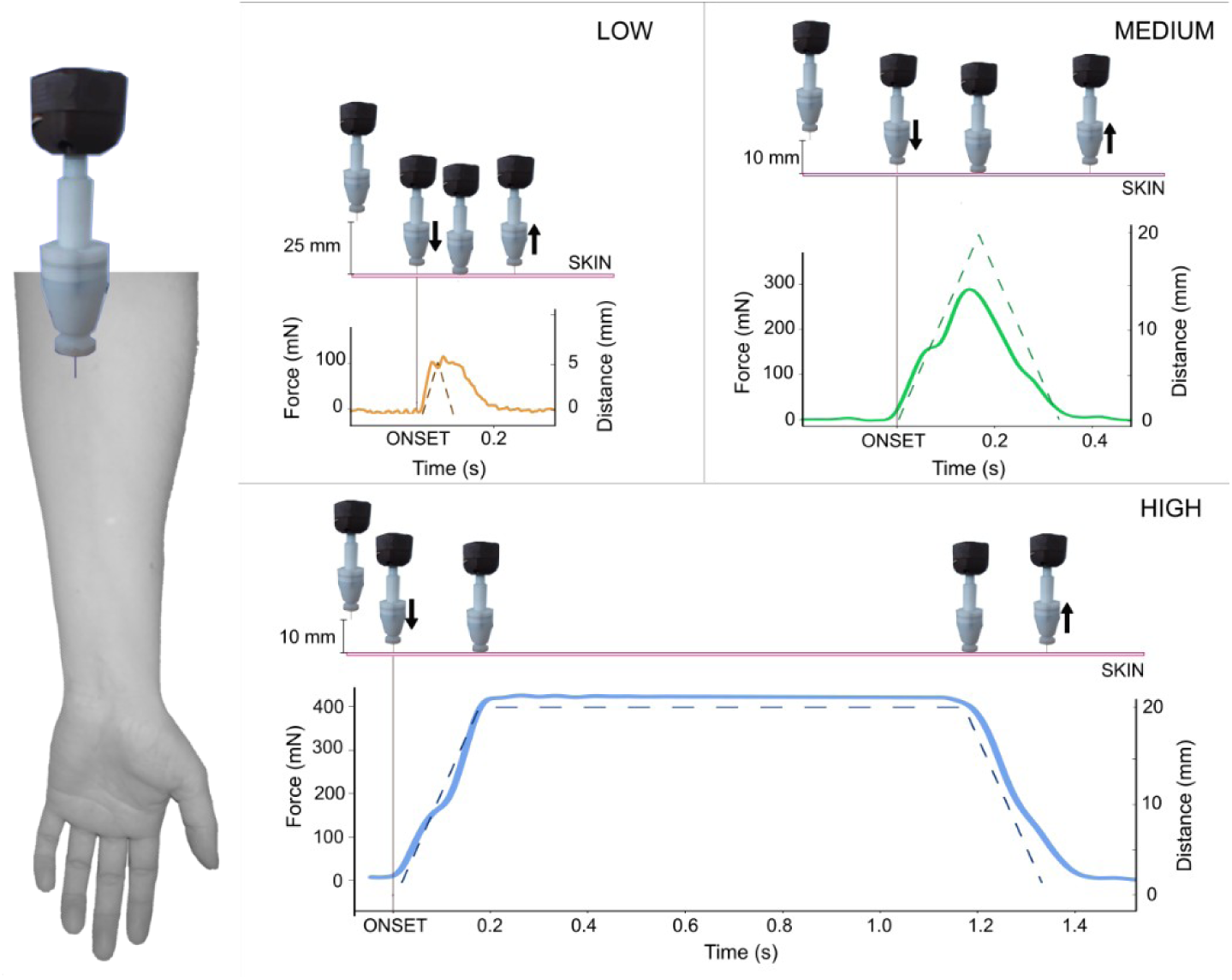
Experimental setup. (Left): Stimulated area on the volar side of the right arm. (Right): The solid lines represent the average force exerted for three stimulation levels: low, medium, and high. Dashed lines indicate the theoretical displacement of the stimulation tip over time, starting from the moment of skin contact (onset). The initial position corresponds to the tip’s distance from the skin before contact.

### EEG and force recordings

Continuous EEG and force signals were recorded together using a wireless EEG amplifier (Cyton, OpenBCI, USA) from seven electrodes mounted in an elastic cap. The electrode positions followed the international 10 *–* 20 system (Fp1, F3, Fz, F4, C3, Cz, C4), with A1 and A2 as reference and ground respectively. EEG and force signals were amplified and digitized using a sampling rate of 250 Hz.

### Perceived intensity ratings and stimulus quality

Volunteers were asked to rate the perceived pinprick intensity after each stimulus using a numerical rating scale (NRS), ranging from 0 (no perception) to 100 (feeling as if the skin was pierced by a needle). Following each block, the quality of perception was assessed using a subset of dimensions of the Spanish Questionnaire based on the McGill Pain Questionnaire (MPQ-SV) (Lázaro et al., 1994). Selected dimensions were *Localization I*, *Punctuate pressure*, *Tactile sensitivity*, and *Evaluative category*.

### Experimental setup

Volunteers sat in a chair with an adjustable arm rest to regulate the arm position for stimulation. To reduce artefacts given by muscular and ocular movements, they were instructed to keep their eyes open, fixate a specific point, and remain relaxed during testing. During the stimulation blocks, white noise via headphones was used to mask the sound generated by the pinprick stimulator. The volunteers were instructed to focus on the stimulus during testing; participants rated the pinprick ratings on the NRS after each stimulus and filled out the MPQ-SV after each block.

### Data analysis

The stimulus onset was defined as the time point at which the force reached 5% of the maximum peak force for each block and subject. The detected onset was subsequently verified through visual inspection. Mechanical impulse was calculated as the time integral of the average force during the stimulation period using the *trapz* function in Python. Peak force values are reported for low and medium impulse levels; average force values over the stimulation interval are reported for high impulse levels. The EEG signals were analysed using MNE-Python (Gramfort et al., 2013). EEG signals were band-pass filtered between 0.5 and 30 Hz with a zero-phase Butterworth filter. Then, signals were cut into 2.5-s epochs, starting 500 ms before the stimulus onset. EEG epochs were visually inspected, and artifact-contaminated epochs (e.g., with eye blinks) were discarded. Pinprick evoked potentials (PEPs) were computed by averaging all epochs within each subject and stimulation levels. PEP features (peak amplitudes and latencies) were measured at the negative peak (N2 peak) between 0 and 200 ms after the stimulus onset and between 150 and 400 ms for the positive peak (P2 peak). The criteria for peak detection were based on Gousset et al. (2023) and corroborated through visual inspection of individual evoked responses.

### Statistics

Data are expressed as mean ± standard deviation for normally distributed variables and median [interquartile range] for non-normally distributed variables. Statistical analyses were performed using jamovi v. 2.2.28 and Python v. 3.11.5. A non-parametric Friedman test was conducted to establish differences in perceived intensity ratings across stimulation levels. Durbin-Conover tests were employed to perform post-hoc pairwise comparisons. Differences in PEP features (amplitudes and latencies) due to stimulation levels were analysed using a linear mixed model with random intercepts per subject. PEP waveforms were compared using a spatiotemporal cluster permutation test, implemented with the *spatio_temporal_cluster_1samp_test()* function from MNE Python’s stats library. A threshold corresponding to a p-value of 0.05 was applied, and a total of 1000 permutations were used for the multiple comparison correction. Scatterplots were employed to visually explore a potential association between impulse levels and N2-P2 PEP amplitudes. P values smaller than 0.05 were labelled as statistically significant, but results were further interpreted considering effect sizes and minimal experimentally relevant differences as well (Mista et al., 2023; Schober et al., 2018; Sullivan & Feinn, 2012).

## Results

Sample characteristics are reported in Table 1. Of the 27 enrolled participants, force recordings were available for 26 at the low impulse level (1 excluded), 20 at the medium impulse level (7 excluded), and 23 at the high impulse level (4 excluded) due to hardware issues during data acquisition. Furthermore, the average number of trials used to derive the PEPs after artifact rejection was 38 [36.5 *–* 39.5] for low impulse, 39 [37 *−* 40] for medium impulse, and 38 [36.5 *–* 39.5] for high impulse stimulation.

**Table 1:**
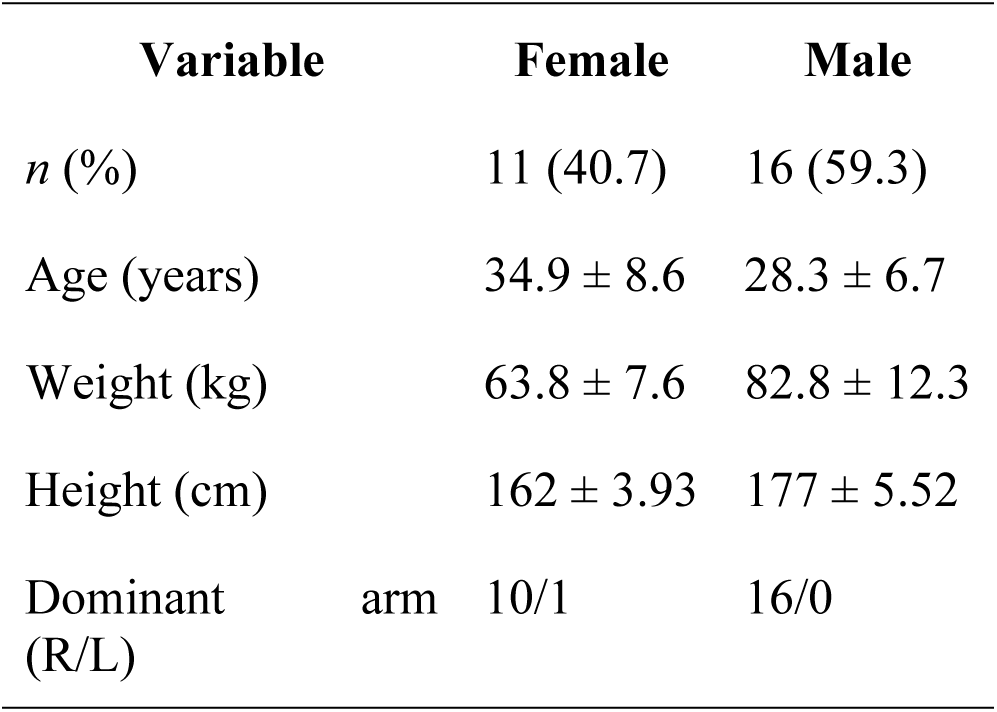
Sample characteristics. Values are expressed as mean ± SD. R, right; L, left.

### Pinprick stimulation

The peak force recorded (mean ± SD) was 144 *±* 57.8 mN, 298 *±* 99.6 mN, and419 *±* 133 mN for low, medium and high impulse stimulation, respectively. The corresponding impulse values were 13.5 *±* 9.26 mN·s, 56.7 *±* 25.6 mN·s, and 505.7 *±* 313.5 mN·s, respectively. The temporal profile for each stimulation level is depicted in Figure 2.

**FIGURE 2.**
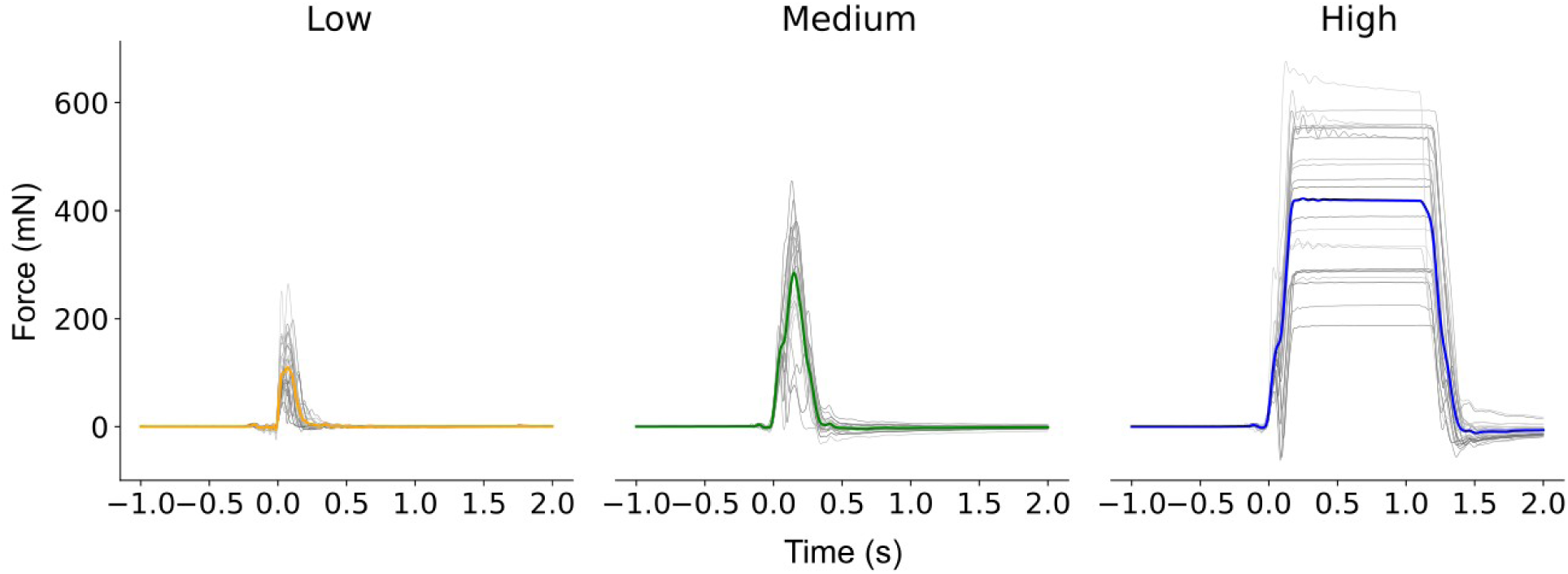
Time courses of force recordings for each impulse level. Gray traces represent the subject-wise average. Coloured traces indicate the grand-average force across all subjects for low (orange, *n*=26), medium (green, *n*=20), and high (blue, *n*=23) impulse stimulation.

### Perceived intensity ratings and quality descriptors

Perceived intensity ratings for each stimulus are shown in Fig. 3. The ratings were 2.5 [1.14 *–* 8.15] for low, 16.4 [ 8.86 *–* 26.9] for medium, and 28.5 [10.2 *–* 41.4] for high impulse stimuli. A main effect of stimulation level was observed ( *χ*^2^ (2)=40.2 *, p*<0.001). Post hoc tests revealed significant differences between all pairwise comparisons: low vs. medium impulse ( *T* =8.72 *, p*< 0.001), low vs. high impulse (*T* =11.9 *, p*<0.001), and medium vs. high impulse ( *T* =3.17 *, p*=0.003).

**FIGURE 3.**
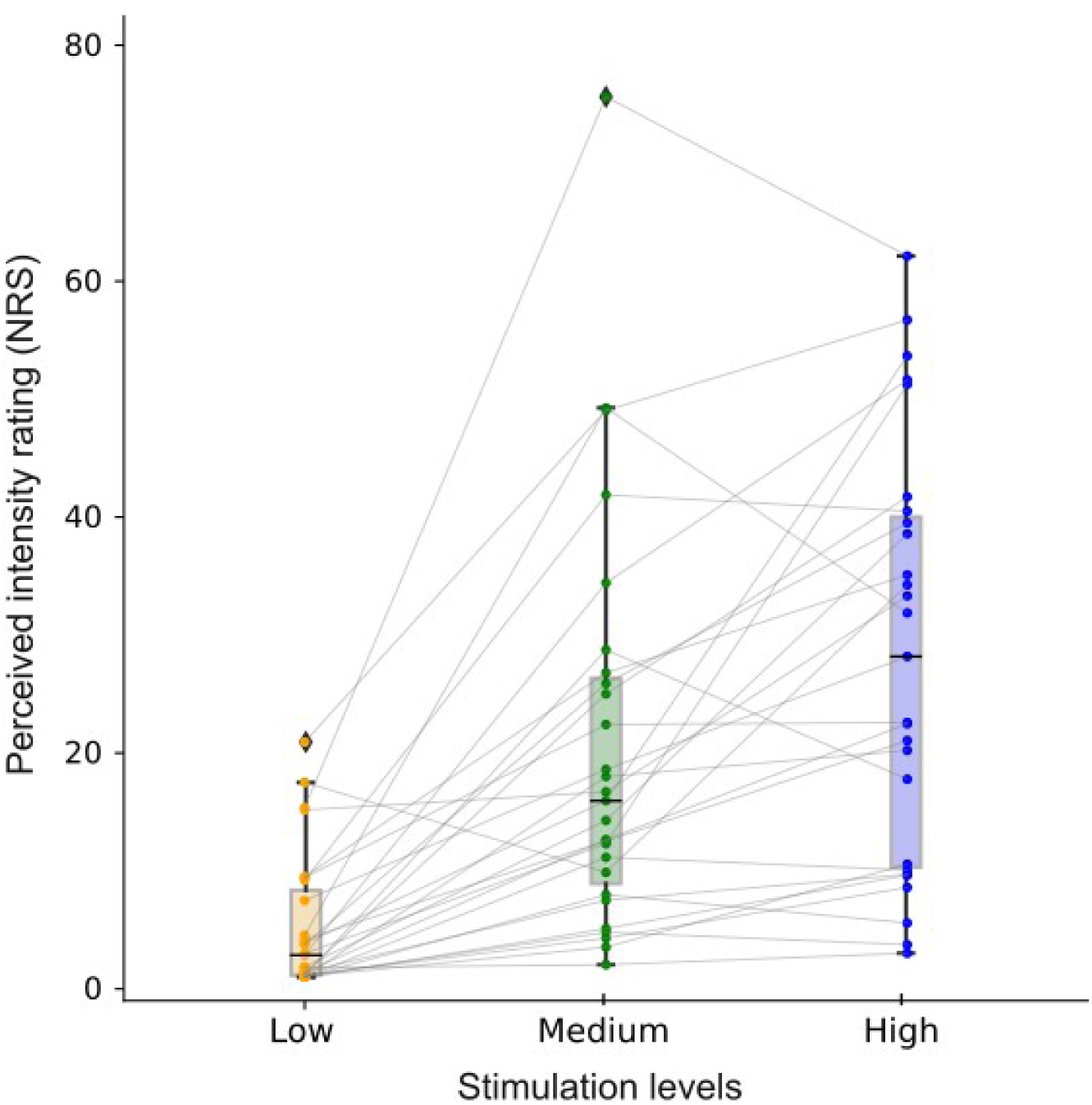
Comparison of perceived intensity ratings across stimulation levels. Individual points represent each volunteer’s median ratings (*n*=27). Grey lines depict repeated measures across participants. Boxes show the median and interquartile range (25*^th^ −* 75*^th^* percentiles), while whiskers represent the 5*^th^*and 95*^th^*percentiles of the ratings across all subjects.

The frequency of descriptors used to characterize pain quality is presented in Table 3. The most frequently reported descriptors were “well-defined” and “pricking”, and these remained consistent across all stimulation levels.

**TABLE 3.** Frequency of selection for descriptors of pain quality of the McGill Pain-Spanish Version Questionnaire for each impulse level.

| Subscale | Descriptor | Low<br>(N \| %) |  | Medium<br>(N \| %) |  | High<br>(N \| %) |  |
| --- | --- | --- | --- | --- | --- | --- | --- |
| Localization I | Imprecise | 5 | 19,23 | 1 | 3,85 | 5 | 18,52 |
|  | Well-defined | 21 | 80,77 | 23 | 88,46 | 18 | 66,67 |
|  | Extensive | - | - | 2 | 7,69 | 3 | 11,11 |
|  | None of the above | - | - | - | - | 1 | 3,70 |
| Punctuate pressure | Pricking | 15 | 57,69 | 14 | 53,85 | 9 | 33,33 |
|  | Boring | - | - | 3 | 11,54 | 5 | 18,52 |
|  | Drilling | - | - | - | - | 1 | 3,70 |
|  | Stabbing | 1 | 3,85 | 7 | 26,92 | 8 | 29,63 |
|  | Lancinating | - | - | 1 | 3,85 | 3 | 11,11 |
|  | None of the above | 10 | 38,46 | 1 | 3,85 | 1 | 3,70 |
| Tactile sensitivity | Rubbing | 14 | 53,85 | 3 | 11,54 | 1 | 3,70 |
|  | Tingling | 3 | 11,54 | 4 | 15,38 | 3 | 11,11 |
|  | Scratching | - | - | 1 | 3,85 | - | - |
|  | Scraping | - | - | 2 | 7,69 | 4 | 14,81 |
|  | Stinging | - | - | 2 | 7,69 | - | - |
|  | Itchy | 5 | 19,23 | 7 | 26,92 | 8 | 29,63 |
|  | None of the above | 4 | 15,38 | 7 | 26,92 | 11 | 40,74 |
| Evaluative category | Weak | 23 | 88,46 | 12 | 46,15 | 4 | 14,81 |
|  | Tolerable | 3 | 11,54 | 11 | 42,31 | 15 | 55,56 |
|  | Intense | - | - | 2 | 7,69 | 7 | 25,93 |
|  | Terribly annoying | - | - | - | - | - | - |
|  | None of the above | - | - | 1 | 3,85 | 1 | 3,70 |

\*Note: The descriptors from the English version of the McGill Pain Questionnaire and the McGill Pain Questionnaire-Spanish Version (MPQ-SV) are dissimilar, we decided to report the translation and not the equivalent descriptors as in the original version.
Shaded values indicate the highest percentage within each descriptor category and impulse level.

### Pinprick evoked potentials

Clusters for the differences between stimulation levels are represented in Fig. 4. Within the centrally selected region of interest, the comparison between high and low impulse revealed significant differences in both early ( *p*=0.048) and late post-stimulus periods ( *p*=0.001). For the high vs. medium impulse comparison, a significant difference was observed primarily in the late window ( *p*=0.001). Finally, the low vs. medium impulse comparison revealed a significant early difference ( *p*=0.038). These results suggest temporally distinct cortical responses to different levels of mechanical stimulation.

**FIGURE 4.**
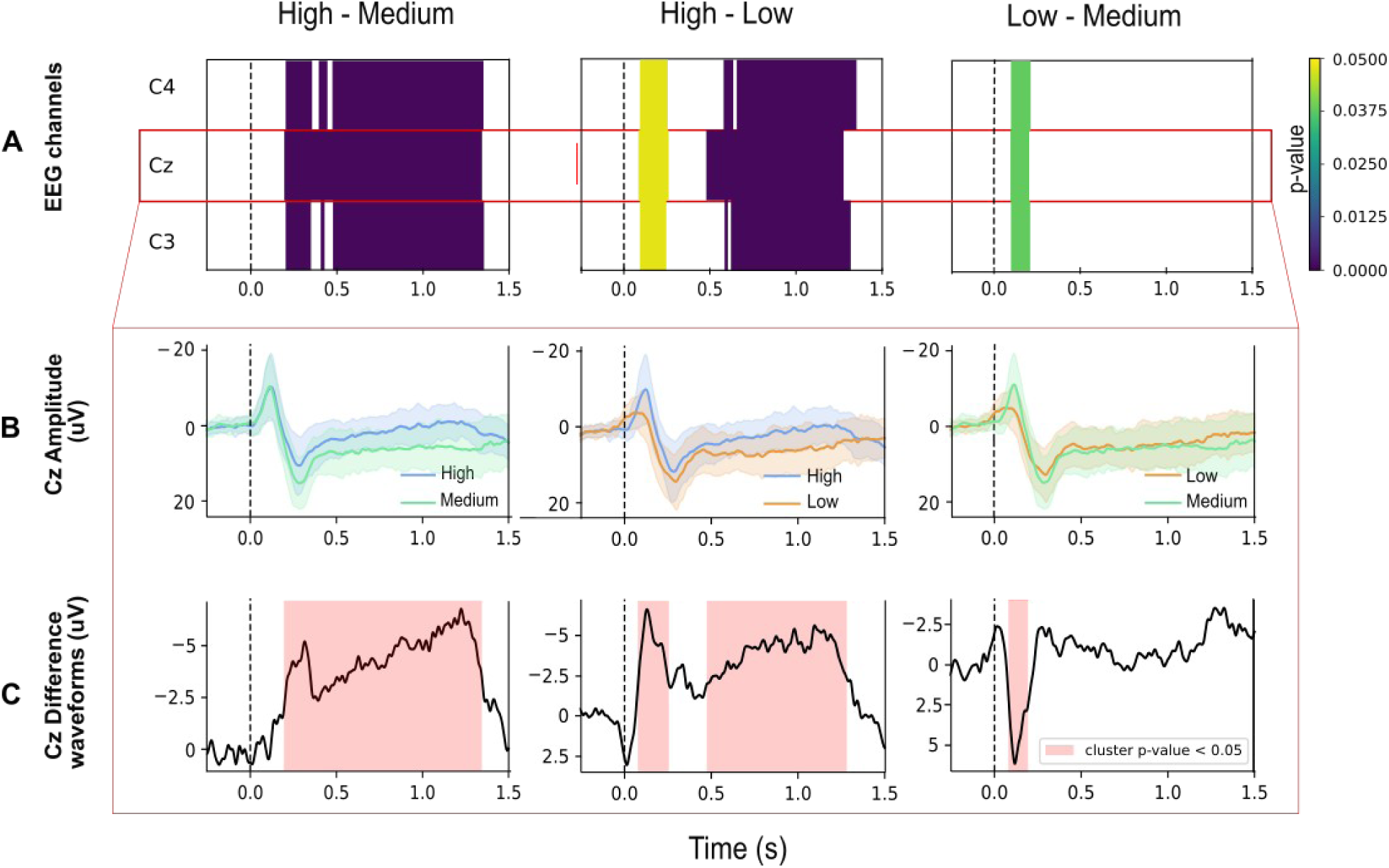
A) Average pinprick evoked potentials for different impulse levels. (Top) Clusters for the different stimulation levels comparison in the channels C3, Cz and C4. The coloured regions represent the found clusters. **B)** Group-level grand-average evoked potentials measured from Cz against A1A2. Shading indicates the standard deviation. **C)** Difference waveforms obtained by subtracting one level from another. Vertical dashed lines indicate the stimulus onset.

The shortest latencies of N2 ( *F*_2,52_=12.9 *, p*< 0.001) and P2 ( *F*_2,52_=2.30 *, p*=0.110) were observed for low impulse stimulation (Fig. 5 left), where the N2 peak occurred at 75.3 *±* 44.3 ms and the P2 peak at 264.3 *±* 49 ms. For medium impulse stimulation, the N2 latency was on average 104 *±* 28.9 ms, with the P2 peak at 284 *±* 33.6 ms. Furthermore, the N2 peak had an average latency of 114.5 *±* 29.8ms while the P2 peak occurred at 277.5 *±*39.2 ms for high impulse stimulation. Post hoc comparisons revealed that N2 latency was longer for high impulse compared to low impulse stimulation ( *p_holm_*< 0.001)and for medium impulse compared to low impulse stimulation ( *p_holm_*=0.001). No difference was observed between high and medium impulse stimulation ( *p_holm_*=0.195). Finally, no differences were observed in P2 latencies across stimulation levels.

**FIGURE 5.**
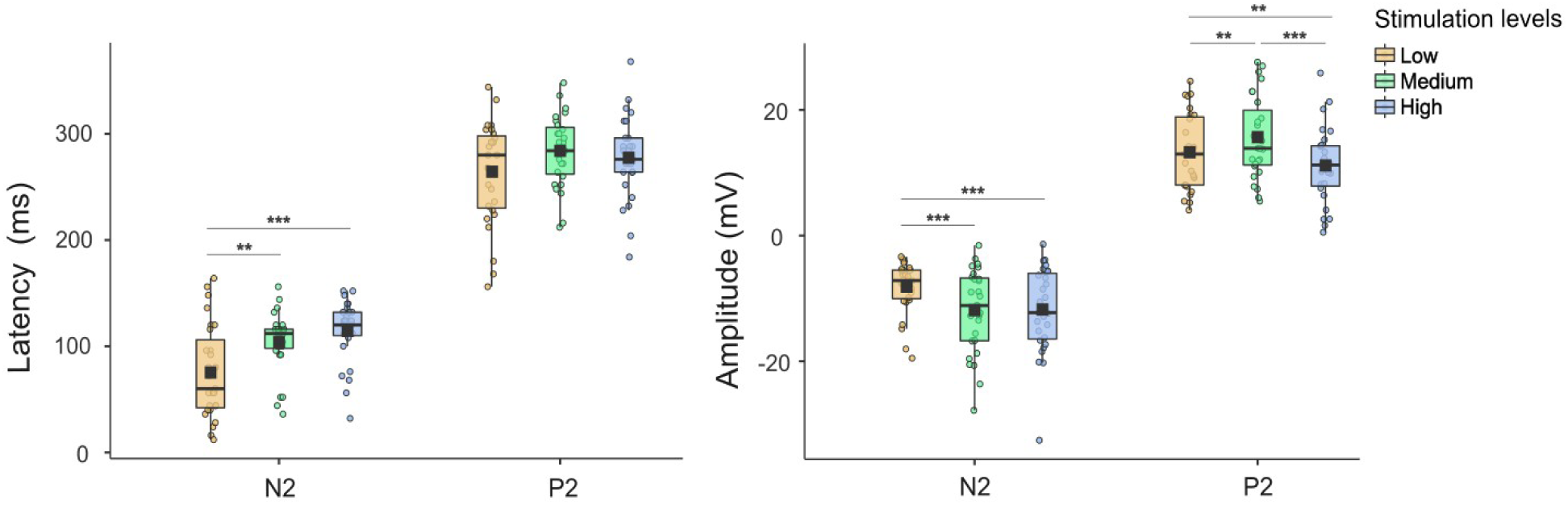
N2 and P2 latencies (left) and amplitudes (right) of pinprick evoked potentials recorded at Cz, across stimulation levels. Individual points show values for each subject (*n*=27).

In terms of amplitudes, the largest N2 amplitude ( *F*_2,52_=11.3 *, p*<0.001) was observed for medium stimulation (*−*11.97 *±*6.78 µV), followed closely by high impulse stimulation µV). In contrast, low impulse stimulation evoked a smaller N2 amplitude of *−* 8.23*±* 4.18 µV (Fig. 5 right). Post hoc comparisons revealed significantly larger N2 amplitudes for both medium versus low ( *p_holm_*< 0.001) and high versus low stimulation ( *p_holm_* < 0.001), with no difference between medium and high ( *p_holm_*=0.900). Regarding P2 amplitudes, the largest response ( *F*_2,52_=13.2 *, p*<0.001) was again observed for medium stimulation (15.58 *±*6.53 µV), followed by low (13.17 *±* 6.11 µV) and high stimulation (11.06 *±* 6.08 µV). Post hoc comparisons showed significant differences between all stimulation levels: medium versus low ( *p_holm_* =0.017), high versus low ( *p_holm_* =0.020), and high versus medium ( *p_holm_*< 0.001).

### Association between stimulation levels and their perceptual and electrophysiological responses

On an exploratory basis, we examined the relationship between stimulation impulse (in mN·s) and both the N2–P2 peak-to-peak amplitude (in µV) and the perceived intensity ratings (on a 0 *–* 100 NRS scale) across stimulation levels. Fig. 6 shows individual averages per condition for all participants. Although visual inspection suggests substantial variability across individuals, no consistent trend was observed within or across stimulation levels. Therefore, no inferential statistical tests were conducted.

**FIGURE 6.**
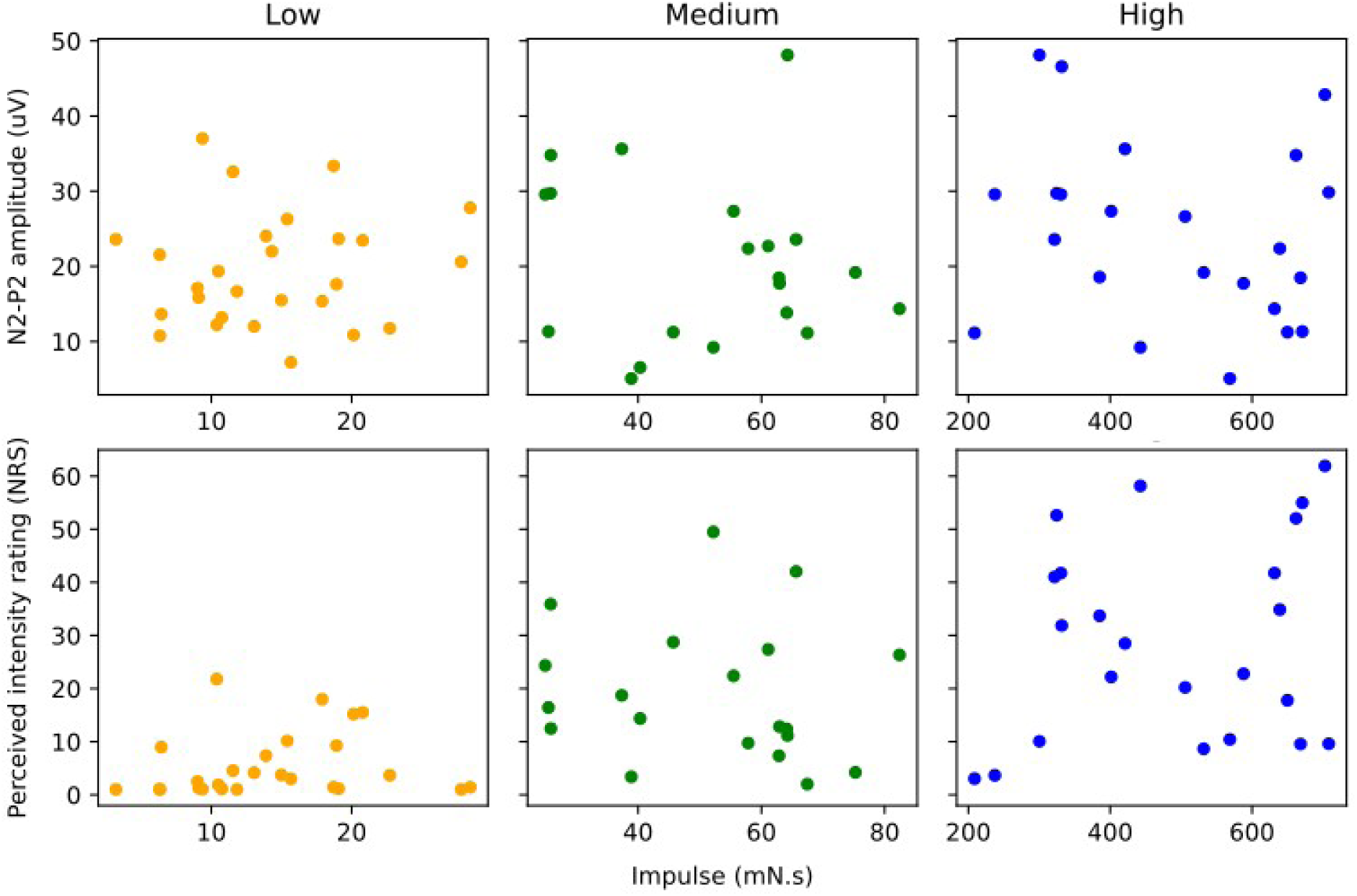
Scatter plots showing the relationship between mechanical impulse (mN·s) and two outcome measures across stimulation levels: top row shows N2–P2 peak-to-peak amplitude (µV), bottom row shows perceived intensity rating (NRS). Each dot represents the average response of an individual participant.

## Discussion

This study characterized the cortical responses and subjective perceptions elicited by pinprick stimuli at different impulse levels using a high-speed robot-controlled stimulator. Higher impulse stimulation levels (i.e., larger forces and longer duration) were associated with larger perceived intensity ratings. Stimulation levels also influenced the PEPs, showing differences in waveforms and characteristic features (latencies and amplitudes) across levels.

### Skin biomechanics and subjective response

Nociceptors are activated by mechanical stress exerted on the skin. Their density is a relevant parameter, as they are not uniformly distributed across the body. Previous studies have demonstrated a proximal-distal gradient in epidermal nerve fibre density, with approximately 60% higher density in the upper thigh compared to the distal leg (McArthur et al., 1998). This gradient has been linked to differences in threshold intensity for pinprick sensation using laser stimulation, and to the proportion of pinprick-evoked potentials elicited when stimulating different dermatomes (Agostino et al., 2000; Rosner et al., 2020). However, despite these regional variations, nerve fibre density within small areas (such as the thigh or the distal leg) exhibits low intra-site variability, suggesting minimal variation within localized regions (McArthur et al., 1998). Likewise, comparable densities have been reported for bilateral biopsies taken from equivalent locations (Lauria et al., 2015). These findings suggest that nerve fibre density is unlikely to account for the variability in electrophysiological and perceptual responses observed in this study, since pinprick stimulation was consistently applied to a small area of the volar forearm. Nonetheless, it may still play a role when comparing results across distinct body regions.

Given this, inter-subject variability (most likely due to differences in the mechanical properties of the skin) and intra-subject variability (linked to subtle variations in stimulus location or habituation to repeated stimulation), can be expected in pinprick experiments. In addition to nociceptor density, the biomechanical properties of the skin and the underlying soft tissue layers (e.g., fat and muscle) are crucial factors in mechanical stimulation (Khalsa et al., 1997). Skin tensile strength and elasticity depend mainly on the dermal collagen and elastic fibre network, while subcutaneous fat provides cushioning (Kolarsick et al., 2011). Variability in these structures, influenced by factors such as age, UV radiation exposure, and hydration levels, has been shown to affect the mechanical properties of the skin. For instance, age-related changes lead to a reduction in collagen content, alterations in collagen fibre organization, and decreased of dermal hydration, all of which further reduce the skin’s mechanical stability and elasticity (Cua et al., 1990). These variations also have a significant effect on PEPs, as the reliability of the cortical responses depends on the body region where stimuli are applied (Rosner et al., 2020). Structural differences among individuals, particularly skin stiffness, likely contributed to the observed differences in both perceived intensity and evoked potentials across participants.

Perceived intensity ratings after pinprick stimulation have also been shown to vary depending on the method of delivery. Robotic stimulation tends to elicit higher ratings than manual application, underscoring the influence of stimulation precision and reproducibility (Iannetti et al., 2013; Rosner et al., 2020; van den Broeke et al., 2020). While this experimental setup does not allow for direct comparison with those studies, the present findings are consistent with the idea that mechanical impulse—combining force and duration—plays a key role in shaping perceived sensation. Participants reported increasingly intense sensations as stimulation impulse increased, reinforcing the relevance of this parameter in nociceptive processing. This consideration may become more relevant for longer stimulus durations, given that the skin deforms over time in response to mechanical stress, in conjunction with underlying soft tissues (Singh & Chanda, 2021). Accordingly, the dynamics of tissue deformation likely contributed to the perceptual and cortical differences observed across stimulation levels in this study.

### Influence of stimulation parameters on pinprick evoked potentials

There is an ongoing debate about the dichotomous classification of mechanical sensory afferents into fibre populations. Aδ and C afferents were prominently associated with conduction of nociception, whereas Aβ afferents were believed to exclusively signal the discriminative aspects of non-nociceptive stimulus (Basbaum et al., 2009; Löken et al., 2009; Wellnitz et al., 2010). However, recent findings challenge this classification. In animal studies, conduction velocities have been shown to substantially overlap across fibre types, and inflammation can increase the conduction speed of nociceptive Aδ fibres, with some reaching velocities comparable to Aβ fibres (Djouhri & Lawson, 2001). Furthermore, circumferential high-threshold mechanoreceptors (Circ-HTMRs) have displayed morphological characteristics of touch receptors, yet functioning as nociceptors (Ghitani et al., 2017). Likewise, studies with humans have identified A-fibre high-threshold mechanoreceptors (A-HTMRs) with conduction velocities comparable to Aβ fibres, capable of encoding noxious mechanical stimuli (Nagi et al., 2019) These findings question the strict dichotomy between nociceptive and non-nociceptive fibres and underscore the utility of mechanical stimulation as a valuable tool for studying pain physiology.

In line with previous reports, the evoked responses in this study displayed a characteristic N2-P2 morphology, and the latencies of the N2 component were consistent with earlier studies using pinprick stimulation (Iannetti et al., 2013; Rosner et al., 2020; van den Broeke et al., 2020). Although Iannetti et al. (2013) used manual stimulation, their parameters are comparable to the medium impulse level in terms of force and duration, yielding a N2 latency (corrected for trigger delay) of 111*±* 7.8 ms, similar to the latencies obtained in this study (104 *±* 28.9 ms). In contrast, the robot-controlled stimulation used by van den Broeke et al. (2020), resulted in longer N2 latencies, which more closely align with the high impulse condition presented here (136 *±* 25 ms vs. 114.5 *±* 28.9 ms, respectively). These differences in N2 latency across studies may be attributed to variations in stimulation parameters. For instance, the stimulation velocity used in the robot-controlled study (33 mm/s) was substantially lower than in this study (125 mm/s), likely contributing to differences in the synchronization of nociceptor activation. Slower stimulation rates tend to reduce synchronization of afferent fibres activation, leading to wider and slower cortical responses.

The low impulse stimulation level showed shorter N2 latency compared to the rest of the impulse levels. The latencies observed for this level are more comparable to the peripheral conduction velocities of low-threshold mechanoreceptors rather than nociceptors. However, we cannot rule out the activation of afferent fibres, particularly Aδ fibres, as some nociceptors may also exhibit short latencies (Nagi et al., 2019). Interestingly, participants predominantly described the perception as pricking, despite latencies consistent with Aβ fibres. The processing of the mechanical stimuli is complex phenomenon, involving a range of peripheral afferents, potentially including C-fibres, Aδ fibres, and low-threshold mechanoreceptors (Slugg et al., 2000, 2004). Finally, data did not show a clear association between the magnitude of stimulation, perceived intensity ratings, and N2-P2 amplitudes. In general, higher stimulation levels yielded higher perceived intensities and larger cortical responses, but considerable overlap and inter-individual variability were present across levels. This suggests that while impulse is a critical factor modulating both subjective and cortical responses, the encoding of stimulus intensity is not entirely explained by a simple linear model. Rather, it reflects a complex integration of peripheral input properties, fibre recruitment, and central processing mechanisms.

### Strengths and limitations

To the best of our knowledge, this study is the first to provide real-time recordings of the force exerted by a pinprick stimulator, and to report the resulting mechanical impulse. These findings show the critical role of individual differences in tissue and skin properties when designing stimulation protocols. Moving forward, a force-controlled system could enhance the precision and consistency of pinprick stimuli by ensuring the reliability of the applied force and measuring the distance covered by the stimulator. EEG has a low signal to noise ratio and therefore the robotic system may introduce some electrical noise into the recordings. However, this was minimized in the experimental setup and accounted for in the interpretation of the findings.

## Conclusion

Cortical and subjective responses to pinprick stimulation are influenced by the stimulus impulse. Studies investigating mechanical punctuate stimulation and evoked responses should consider both inter-subject variations, driven by differences in skin mechanical properties, and intra-subject variability, influenced by stimulus location.

## Data Availability

All data produced in the present study are available upon reasonable request to the authors

## Data Availability

Data is available under request to authors

## Acknowledgments

We would like to express our sincere gratitude to Ariel Dario Kinderknech and Erick Arnolds for their crucial contributions to the development of the pinprick stimulator. We thank Flavio Lovatto for his assistance in the data collection process.

